# Death by Round Numbers: Glass-Box Machine Learning Uncovers Biases in Medical Practice

**DOI:** 10.1101/2022.04.30.22274520

**Authors:** Benjamin J. Lengerich, Rich Caruana, Mark E. Nunnally, Manolis Kellis

## Abstract

Real-world evidence is confounded by treatments, so data-driven systems can learn to recapitulate biases that influenced treatment decisions. This confounding presents a challenge: uninterpretable black-box systems can put patients at risk by confusing treatment benefits with intrinsic risk, but also an opportunity: interpretable “glass-box” models can improve medical practice by highlighting unexpected patterns which suggest biases in medical practice. We propose a glass-box model that enables clinical experts to find unexpected changes in patient mortality risk. By applying this model to four datasets, we identify two characteristic types of biases: (1) discontinuities where sharp treatment thresholds produce step-function changes in risk near clinically-important round-number cutoffs, and (2) counter-causal paradoxes where aggressive treatment produces non-monotone risk curves that contradict underlying causal risk by lowering the risk of treated patients below that of healthier, but untreated, patients. While these effects are learned by all accurate models, they are only revealed by interpretable models. We show that because these effects are the result of clinical practice rather than statistical aberration, they are pervasive even in large, canonical datasets. Finally, we apply this method to uncover opportunities for improvements in clinical practice, including 8000 excess deaths per year in the US, where paradoxically, patients with moderately-elevated serum creatinine have higher mortality risk than patients with severely-elevated serum creatinine.

## Introduction

Analyzing real-world evidence (RWE) is difficult because RWE includes observations of patients who may be receiving interventions. Interventions are selected in consideration of patient risk factors, so data-driven risk models can confound patient risk with effects of interventions and produce implausible statistical effects (Figure 1). For example, patients hospitalized with pneumonia have been observed to have *lower* mortality risk if they *also* have asthma [1]. This finding contradicts biological causality since pneumonia is complicated by asthma [2], but it reflects an aspect of medical practice – patients with known risk factors like asthma may request and receive more urgent care.

**Figure 1:**
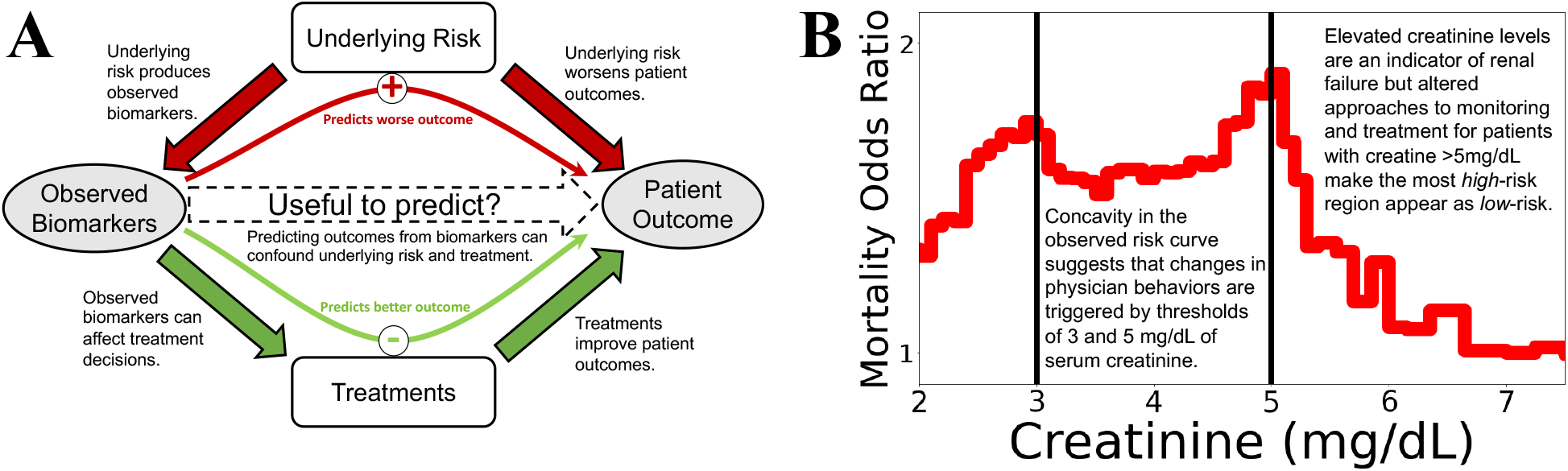
Confounding effects are treacherous to data-driven risk models, but confounding effects that are revealed by glass-box models can be useful by suggesting potential improvements in medicine. **(A)** Underlying “treatment effects” confound risk models. Causal arrows are filled, observed variables are shown in gray ovals and unobserved variables in white boxes. Data-driven analyses often estimate ℙ (Outcome|Biomarker), but this is only a faithful surrogate for ℙ (Outcome|Underlying Risk) if treatments were to have negligible impacts. In reality, treatments (broadly interpreted, including monitoring, therapeutics, diagnostics, and patient behavior) have large impacts on outcomes. When explicitly analyzed for treatment effectiveness, effects of randomly-assigned treatments are desirable as evidence of a proposed treatment being effective, but strong treatment effects confound estimation of risk. To build models which can effectively guide treatment decisions, we require intelligible models and medical domain knowledge to understand if all relevant confounders have been sufficiently corrected. **(B)** An example of strong, but useful, confounding: the mortality risk of pneumonia patients falls with extremely high levels of serum creatinine (which indicates kidney failure), even after correcting for other risk factors in a multivariable predictive model. This counter-causal relationship suggests confounding, and the sharp inflections at the round numbers of 3mg/dL and 5mg/dL (denoted by black vertical lines) suggest that this association is guided by discrete treatment thresholds rather than smooth biomedical risk factors. While this confounding between risk factor and clinical decisions is a challenge for data-driven analysis, it is also an opportunity because the unexpected inflections alert us to possibilities of optimizing treatment.

This example of asthma and pneumonia is doubly problematic in the quest for data-driven medicine: not only would a naíve artificial intelligence (AI) model learn an effect that contradicts medical causality (“asthma is good for pneumonia”), but also this contradiction results in the model predicting lower intrinsic risk for the patients for whom standard treatment would be most beneficial (“pneumonia patients with asthma are low risk and thus don’t need treatment”). In general, data-driven risk models will assign low risk to patients who could be effectively treated. While physicians can compensate for clinical context while interpreting data-driven risks [3, 4], without this contextual grounding, data-driven models faithfully recapitulate the effects (and biases) of clinical practice. This confounding is not an indictment of limited *samples* or subpar *models*: treatment effects often propagate with consistent patterns, so any model that accurately predicts observed outcomes will invariably learn these spurious effects.

Confounding by treatments is also not a problem of limited *features*. While confounding in medical informatics has been widely studied [5, 6] and many computational approaches have been proposed to overcome this challenge [7–9], there are problems with strategies that use only statistical methods to correct confounding. First, many interventions are selected based on risk factors without randomization, so there is limited statistical identifiability to separate the impact of treatments from the impact of risk factors. Second, many influences on treatment decisions are not recorded or observable (e.g. patient urgency). Third, without tools to audit data-driven models, it is difficult to know if all confounding effects have been identified and corrected.

We see the challenge of confounding as an opportunity. When data-driven risk robustly defies biological causality, there may be an opportunity for improving healthcare. For every paradoxically low risk region, there is also a paradoxically high risk region. For example, if risk paradoxically decreases at a threshold, then the risk curve implies at least three conclusions: (1) this threshold is used for guiding treatment decisions, (2) the treatment is effective, and (3) patients just below the threshold could potentially have risk reduced but are not receiving the treatment. Moreover, thresholds often occur at round numbers, suggesting that treatment decisions may be suboptimal and can be improved. By providing interpretable risk curves, transparent (“glass-box”) risk models empower medical experts to detect these paradoxical statistical effects caused by confounding and uncover opportunities for optimizing medical practice. In this way, we can transform the challenge of confounding into an opportunity to improve healthcare.

We apply glass-box machine learning to study four large datasets and find two main types of treatment effects: discontinuous risk frequently seen at round numbers, and paradoxical risk in which successful treatments reduce the risk of higher-risk patients below the risk of untreated lower-risk patients. We first examine idealized treatment effects in simulation, and then examine the pneumonia dataset introduced above to identify characteristic types of treatment effects. Next, we study several versions of the widely-used MIMIC dataset, which has been collected over several decades [10–12], to examine how treatment effects have changed over time. Finally, we estimate the excess mortality attributable to reliance on round-number thresholds and identify areas for improvement with personalized medicine.

## Results

### Threshold-guided clinical practice produces recognizable statistical artifacts

To understand the impacts of confounding from threshold-guided clinical practice, we examine risk curves of simulated treatment benefits and protocols (Figure 2). We study eight simulation cases; for each simulation case, we select one of four treatment benefit curves (flattens risk, limits risk, reduces biomarker, or has constant benefit — the left column in Figure 2) and one of two treatment probability distributions (strict adherence to a biomarker threshold, or loose guidance based on a biomarker threshold — the middle column in Figure 2). These underlying risk and treatment curves combine to produce a population risk observable in RWE — the right column in Figure 2. The shape of the observed population risk (right column) is influenced by the shapes of the underlying true risk curves (left column). When the threshold-based distribution of treatment decisions is mis-aligned from the statistically-optimal threshold, the population risk curve contains characteristic artifacts. For example, strict adherence to a threshold that is too high (Figure 2A, yellow) results in observed population risk that includes excess risk below the treatment threshold, with a sharp, discontinuous drop in risk at the treatment threshold.

**Figure 2:**
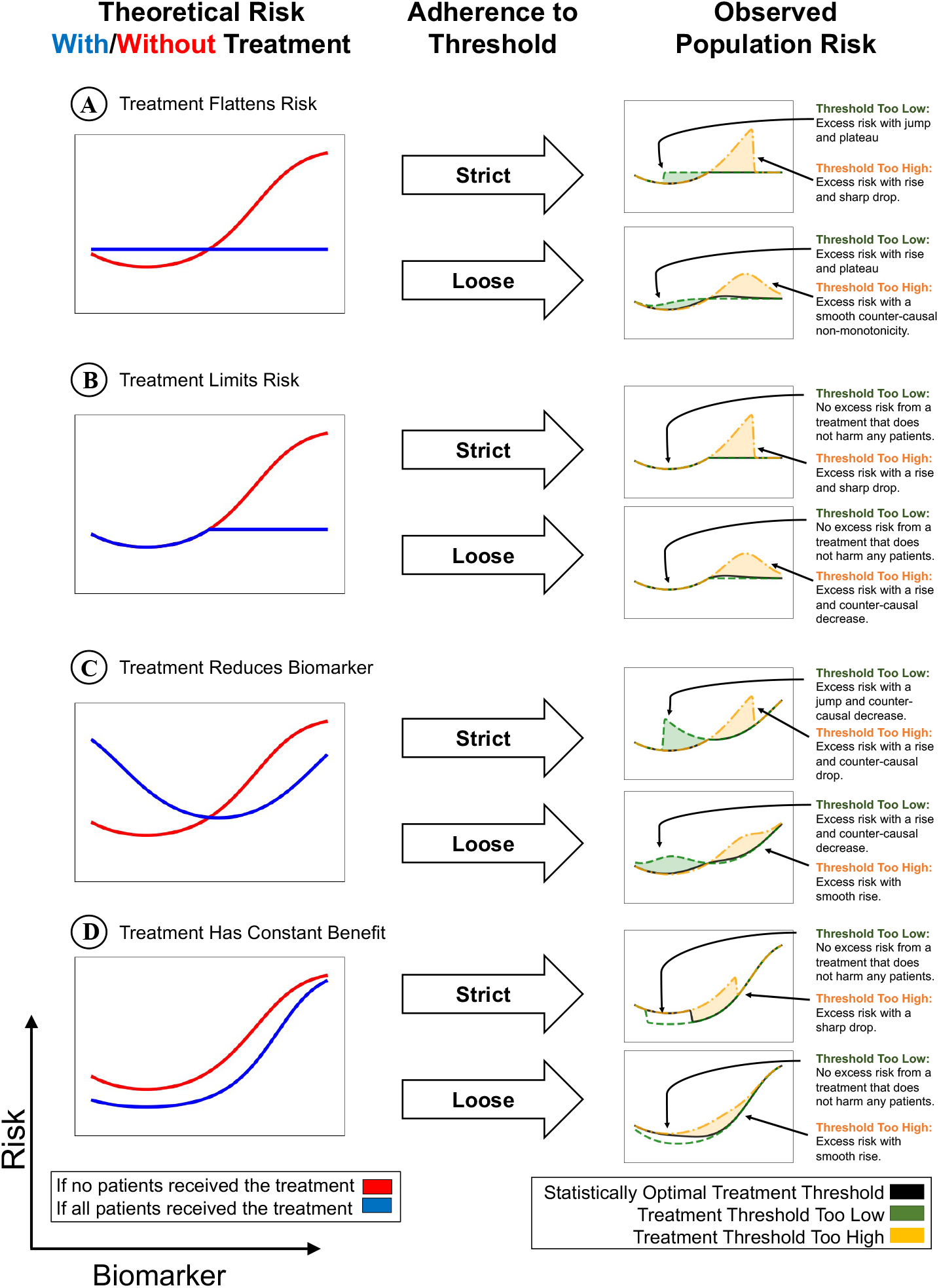
Confounding from treatment effects produces characteristic shapes in population risk. In each row, we depict the underlying intrinsic risk for treated and untreated patients (left), adherence of treatment decisions to a threshold (middle), and observed risk (right). The optimal treatment threshold is the point where untreated risk crosses above treated risk; when treatment protocols are misaligned from optimality, observed risk includes excess risk. **(A)** When treatment induces a constant risk (e.g. surgery), and the treatment threshold is lower than optimal (green), there is a rapid rise in risk followed by a plateau. Conversely, if the treatment threshold is higher than optimal (yellow), there is an overshoot and drop. **(B)** When treatment caps risk (e.g bronchodilators), then no threshold is too low, while a high threshold produces a smooth increase in risk followed by a drop. **(C)** When treatment reduces the biomarker value (e.g. vasodilators reduce blood pressure), a low threshold produces a rapid and/or discontinuous rise in risk followed by a counter-causal non-monotonicity, while a high threshold produces a smooth increase in risk followed by a rapid and/or discontinuous decrease in risk. **(D)** If the treatment induced a constant benefit for all patients (e.g. an idealized treatment), then there is no threshold that is too low, and any threshold produces a counter-causal drop.

These artifacts can be interpreted by combining domain knowledge of treatment benefits to reverse-engineer the effects of clinical decision making from population risk curves and suggest improvements to clinical practice. For example, if we observe the sharp, discontinuous drop in risk that is characteristic of strict adherence to a threshold which is too high, we can suggest a refinement to lower this treatment threshold. Similarly, the slope of the artifact is dictated by the degree of adherence to the threshold: when thresholds are strictly obeyed, the risk curve is discontinuous at the threshold; when thresholds are followed only loosely, the risk curve changes direction in a smooth non-monotonicity. In general, clinical decision distributions that are statistically near-optimal yield smoother and flatter risk curves. As medicine becomes increasingly precise, multi-faceted, AI-supported and personalized, we expect to see risk curves smooth out round-number biases and flatten observed risks — we later discuss an example (Figure 4D) where improvements in clinical decision-making have already smoothed and flattened risk curves.

### A model for systematically identifying statistical artifacts of clinical practice

To identify these recognizable statistical artifacts, we decompose risk into components of individual risk factors and then find regions with discontinuities and non-monotonicities. This requires an accurate and high-resolution model of patient risk that can be decomposed into univariable graphs without sacrificing accuracy. Thus, we trained generalized additive models (GAMs) [13] to estimate mortality risk as additive effects of individual risk factors. The additive model is highly predictive, with accuracies comparable to fully-connected neural networks and XGBoost baselines (Table S1) and provides high-resolution component functions of individual variables which can be analyzed sequentially to uncover discontinuities and non-monotoncities in risk.

#### Automated search

Based on the principles identified in simulation, we propose two methods to automate the search for discontinuities and non-monotonicites in the individual component functions of this trained GAM. First, to search for discontinuities, we compare the probability of observed outcomes under a discontinuity against the probability of the same observed outcomes under a locally-linearized risk curve (Figure S2B). Second, we search for non-monotonicites by applying Bayesian changepoint detection [14] to the non-zero slopes of the risk curve (Figure S2C). These methods provide rankings, effect sizes, and visualizations of identified statistical artifacts, and enable medical experts to interactively identify hidden confounding, biases, and opportunities to improve clinical practice. Details, code, and tools to use these models are available in the Methods section.

### Systematic search reveals pervasive confounding by treatments

We next turn to RWE in large EHR datasets to study the extent and magnitude of hidden confounding by treatments. We study four datasets: the 1989 MedisGroups Comparative Hospital Database (MCHD) pneumonia dataset, [15] and three versions of the Multiparameter Intelligent Monitoring in Intensive Care (MIMIC) dataset [10–12]. By applying the systematic search for artifacts of clinical practice, we find: 6 surprising non-monotonicities in the 19 continuously-valued biomarkers of the Pneumonia dataset, 12 discontinuities and 2 surprising non-monotonicities in the 15 biomarkers of MIMIC-II, 37 discontinuities and 4 surprising non-monotoniciites in the 15 biomarkers of MIMIC-III, and 27 discontinuities and 6 surprising non-monotonicities in the 15 biomarkers of MIMIC-IV. As case studies, here we examine four of the artifacts found in the Pneumonia dataset (Figure 3).

**Figure 3:**
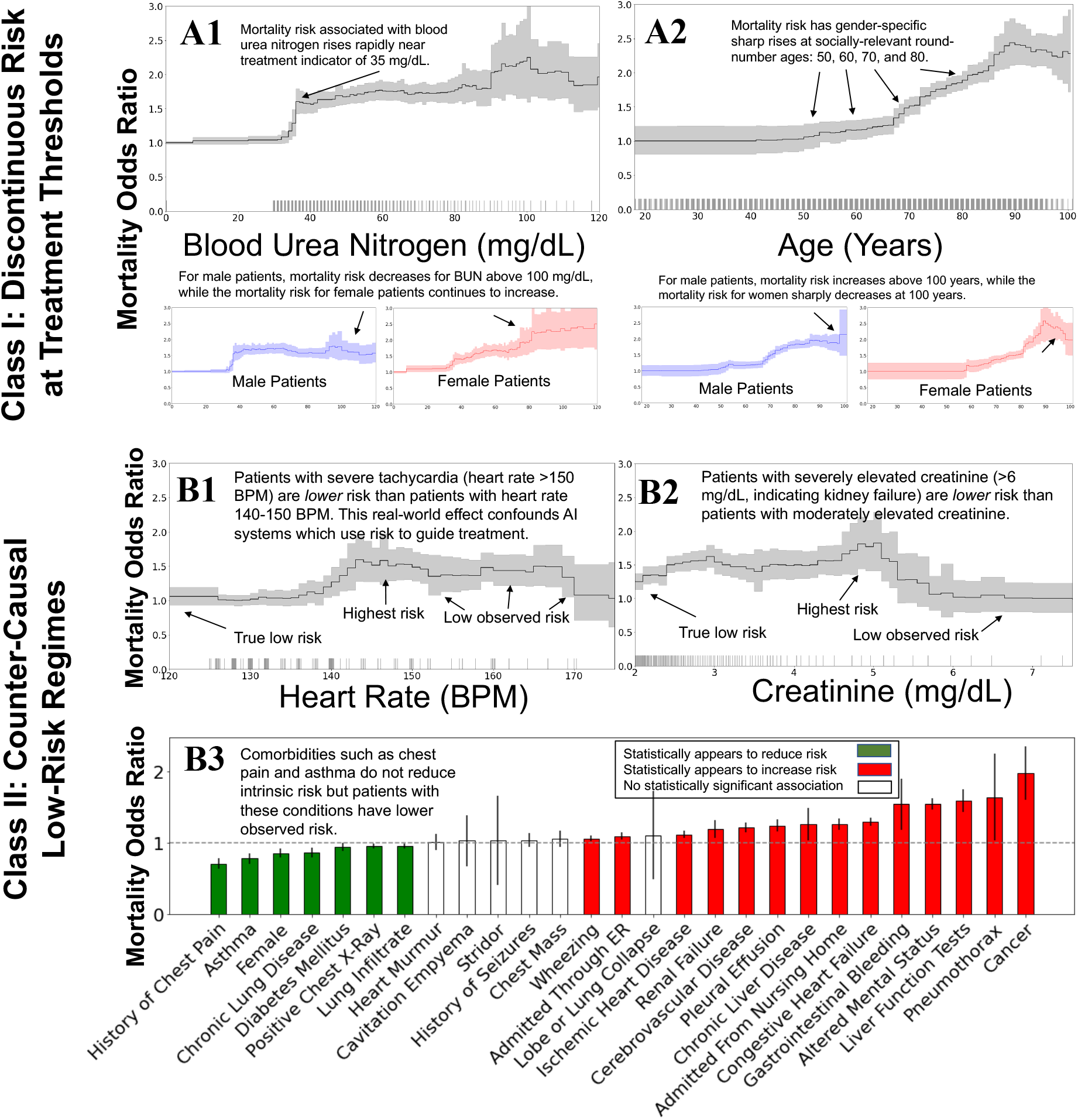
The mortality risks of patients with pneumonia display a number of confounding effects that can be characterized in two broad categories: discontinuities and counter-causal effects. In all plots, we show the effect of each variable on patient mortality risk (with 95% confidence intervals shaded) after correcting for all other observable factors of patient risk using a generalized additive model. Each tick along the horizontal axis denotes 10 observed patients. (**A)** Discontinuities in the observed mortality risk are produced by behavior influenced by discrete round-number thresholds, including blood urea nitrogen above 35 mg/dL and 100 mg/dL (**A1)**, and 10-year increments in age, most notably age 100 for female patients (**A2)**. (**B)** Counter-causal trends in the observed mortality risk curves lead to lab values and comorbidities that are intrinsically high-risk being predicted as low-risk due to effective treatments. This confounding can lead data-driven AI models to prefer lab values that are intrinsically unsafe, including heart rate above 150 BPM (**B1)** and serum creatinine above 5 mg/dL (**B2)**. Finally, chronic comorbidities that increase perceived urgency for treatment, including history of chest pain, asthma, and chronic lung disease, are associated with lower mortality risk (**B3)**, even though these comorbidities increase intrinsic risk.

#### Discontinuities

First, blood urea nitrogen (BUN) has a discontinuous impact on mortality risk (Figure 3A1), with a non-monotone structure that implies different treatments at different BUN levels. Lower levels of BUN are associated with survival; however, there is a rapid rise in mortality risk from 30-40mg/dL, with a plateau from 40-80mg/dL, and possibly a reversal of the trend above 100mg/dL. The amelioration of risk at BUN of 40mg/dL is stronger for male patients than female patients, with the risk for female patients rising continuously from 30-50mg/dL. Finally, the risk associated with elevated BUN continues to rise for female patients while the risk for male patients is relatively flat from 40-120 mg/dL, suggesting an opportunity for improved care of female patients with elevated BUN.

Mortality risk also changes in discontinuous steps with respect to patient age, including increases in mortality risk at several round-number ages: 50, 60, 70, and 80 (Figure 3A2). In addition, there is evidence of a “centenarian effect” for female patients as the mortality risk surprisingly declines approaching age 100. This centenarian effect is the opposite of the so-called “old man’s best friend”[16] effect where pneumonia may be seen as a blessing for elderly patients with potentially terminal and unpleasant comorbidities. In both of these examples (BUN and age), the biases are so strong that they override the smoothness of inherent biological risk to produce discontinuous risk curves with step changes at round numbers and clinically-meaningful thresholds.

#### Counter-Causal Non-Monotonicities

Treatments also produce counter-casual effects where regions of low-risk are produced by aggressive treatment rather than low intrinsic biological risk. For example, elevated heart rate above 145 beats per minute (BPM) (Figure 3B1) and elevated serum creatinine above 5mg/dL (Figure 3B2) are both associated with increased survival (lower mortality risk) of pneumonia patients. These statistical effects (each measured after correcting for all other patient risk factors recorded in the dataset) contradict medical causality: heart rate above 145 BPM corresponds to severe tachycardia while serum creatinine above 5mg/dL indicates advanced renal dysfunction [17], suggesting that intervening factors including aggressive treatment were given to these patients who would otherwise be at extreme risk absent treatment. Unfortunately, purely data-driven AI protocols likely would de-prioritize care for patients with tachycardia and elevated creatinine, and may even use these extreme regions as targets toward which patients should be guided via therapy!

Chronic comorbidities also produce paradoxical statistical effects. For example, history of chest pain, asthma, and chronic lung disease *reduce* the probability of mortality in pneumonia patients by more than 30%, 20%, and 18%, respectively (Figure 3B3); these effects are robustly estimated and not sensitive to the model class or algorithm used to estimate the effects. These effects contradict medical intuition of causality and suggest that risk models trained on these real-world datasets could systematically underestimate mortality risk for patients with prior comorbidities. This systematic underestimation may occur because patients are acutely aware of their own respiratory distress, and their previous experiences, diagnoses and access to healthcare will influence how they react as new problems arise; similarly, clinicians may react differently to patients with different medical histories. As a result, the observed risk is a combination of underlying risk, perceived urgency by both the patient and the clinician, and the impact of treatments. When developing data-driven treatment protocols, we must take care to not disadvantage patients with comorbidities that may have warranted aggressive treatment in the training data. For both lab tests and comorbidities, it is important to identify these counter-causal regions of low risk that are produced by aggressive treatments because these are places where AI models that do not understand the causal impact of effective treatments would mistakenly predict patients to be at low risk, possibly denying them the care needed (and routinely provided) to reduce the risk.

### Clinical practice has improved over years of refinement

To study how clinical practice and treatment effects have changed over years of protocol refinement, we examine three versions of the MIMIC dataset [10–12] that have been used by thousands of researchers to train statistical models of mortality risk. These datasets are high-quality snapshots that were collected over three decades of intensive care units (ICU), providing a large-scale view of medical practice and outcomes over many years. We find that while the negative impact of these treatment effects has decreased over the years, counter-causal and threshold-driven effects are still strong and robustly estimated by data-driven risk models. Finally, we identify treatments recorded in MIMIC-IV [12] that could de-confound the effects of treatment from the underlying biological risk. Overall, we find that the statistical artifacts of clinical practice are associated with observable treatment patterns, typically based at clinically-meaningful and round-number thresholds, and that while these effects have diminished over time, they are still routinely and robustly identified to impact patient mortality risk. Here, we study four examples of these effects.

#### Ex 1: Counter-causal effects of serum creatinine have strengthened

Elevated levels of serum creatinine indicate kidney dysfunction[18]; however, this biomarker is not strongly associated with mortality at extremely high values (Figure 4A). Instead, the highest risk (after correcting for all other patient risk factors) is observed for patients with moderate serum creatinine in the range 3-5mg/dL. The plateau in mortality rate at 3mg/dL is associated with a strongly increased probability of treatment with intermittent hemodialysis (Figure 4A2). Similarly, the prevalence of continuous renal replacement therapy (CRRT) begins to increase at a creatinine level of 3mg/dL and plateaus at 4mg/dL (Figure 4A3). At a serum creatinine level of 5 mg/dL, the prevalence of intermittent hemodialysis begins to decrease while the prevalence of CRRT correspondingly increases, and the mortality rate decreases. These patterns suggest a possibility that the treatments and extra monitoring for kidney dysfunction reduce patient mortality even below the risk level of patients with healthy levels of creatinine. One serious concern is that AI models that seek to minimize mortality probability might underestimate the danger of unhealthy levels of serum creatinine and advise withholding treatment from these patients.

**Figure 4:**
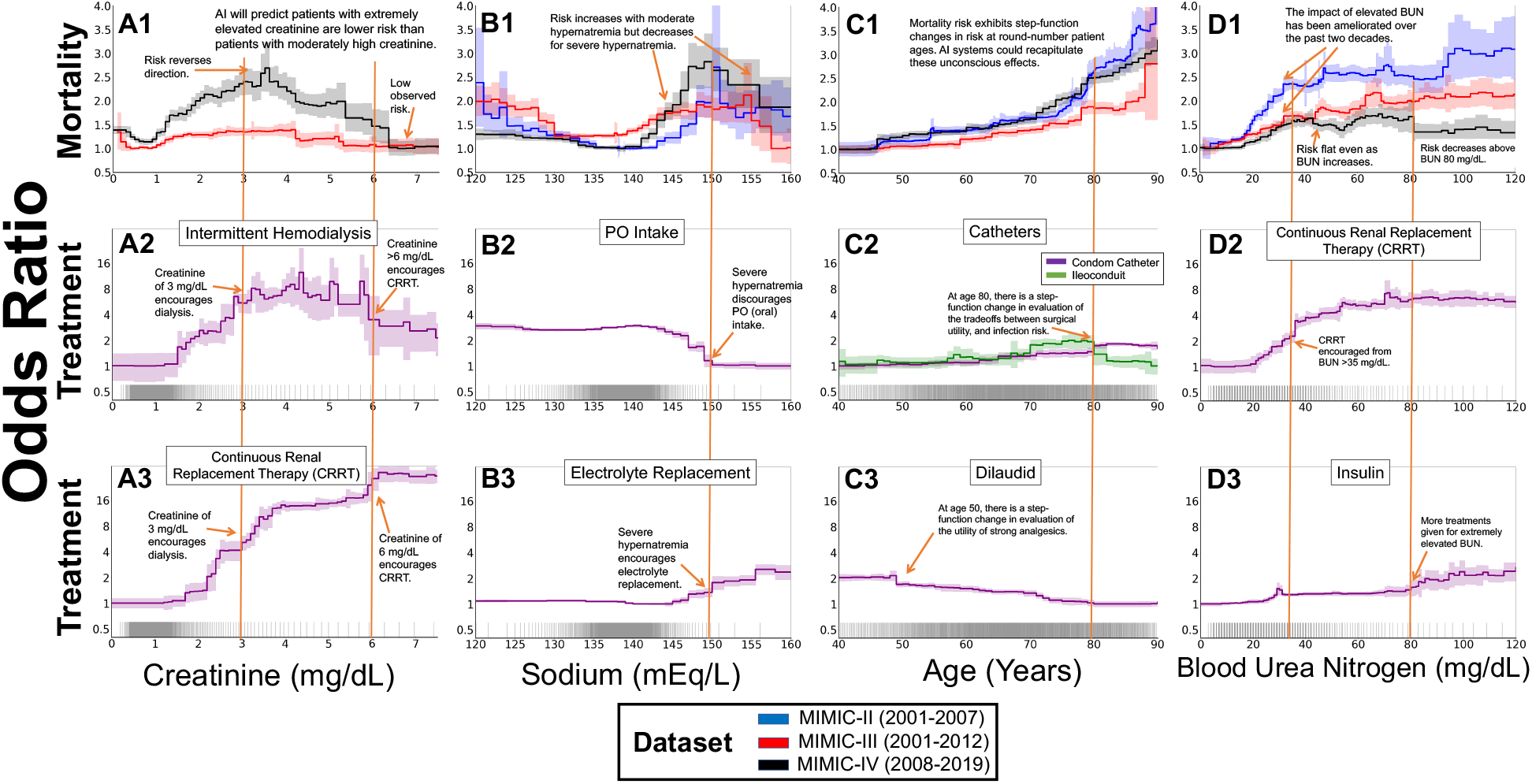
The effects of clinical practice are visible in mortality risk curves for patients in intensive care units, and have changed over time. The three datasets (MIMIC-II, MIMIC-III, and MIMIC-IV) span three decades of intensive care at a single hospital system, allowing comparisons of the effects of clinical practice that have improved over time. The top row shows mortality risk associated with each risk factor after correcting for all other observable factors of patient risk using a generalized additive model (with 95% confidence intervals shaded). The bottom two rows show the treatment propensity associated with each risk factor in MIMIC-IV (with 95% confidence intervals shaded). Ticks along the horizontal axes each denote 10 patients. **(A)** The mortality risk attributable to serum creatinine plateaus at a creatinine level that is only moderately elevated **(A1)**. For extremely elevated creatinine, there is a counter-causal decrease associated with treatment with intermittent hemodialysis **(A2)** or continuous renal replacement therapy **(A3). (B)** The mortality risk attributable to serum sodium decreases at 150 mEq/L **(B1)**, associated with a decreased usage of PO intake (i.e. eating/drinking by mouth, **(B2)**) and an increase in electrolyte replacement (Dextrose 5%, Free Water, Potassium Phosphate, Sterile Water, Sodium Chloride 0.45%)**(B3). (C)** The mortality risk attributable to age increases with discontinuous impacts at 5-year intervals **(C1)**, with a changes in catheterization frequency at age **(C2)** and analgesic prescriptions at 50 **(C3)** demonstrating changing value judgements influencing clinical care as patients progress past round-number ages. The mortality risk attributable to elevated blood urea nitrogen **(D)** exhibits multiple peaks and valleys **(D1)**, with changes in slope linked to propensity for CRRT **(D2)** and insulin prescription **(D3)**. All of these analyses were calculated using a model that is agnostic to round numbers, so the recovery of these effects indicate that the observational data are confounded by real-world treatment decisions influenced by round-number thresholds. These discontinuities suggest opportunities to fine-tune treatment decisions.

#### Ex 2: Counter-causal effects of serum sodium have endured

As expected from medical intuition regarding hydration, the healthy range of 135-145 mEq/L serum sodium is associated with low mortality risk (Figure 4B1). On either side of this healthy range, the probability of mortality increases, rising rapidly for hyponatremia below 135 mEq/L and hypernatremia above 145 mEq/L. However, this medical intuition is broken at 150 mEq/L: the probability of mortality declines for extremely elevated serum sodium. This threshold of 150 mEq/L is an indicator of moderate hypernatremia [19], and is associated with increased attention from care providers, reduced oral (PO) food intake (Figure 4B2) and increased electrolyte and water replacements (Figure 4B3). As a result of aggressive treatment of severe hypernatremia and under-treatment for mild hypernatremia [20], the probability of mortality is *lower* for patients with serum sodium above 155 mEq/L than for patients with serum sodium of 145 mEq/L. This effect is robustly supported by statistical evidence and is clinically meaningful — any accurate model trained on these datasets has learned that severe hypernatremia is lower risk than moderate hypernatremia, so applying these data-driven models in clinical practice could divert treatments away from the group of patients who would benefit *most* from treatment.

#### Ex 3: Discontinuous effects of age have weakened

Patient age exhibits discontinuous impact on mortality rates (Figure 4C1). While the impact of each particular round-number age has changed over the years (possibly due to increases in life expectancy and social changes such as in retirement), age 80 confers increased risk in all 3 three datasets. This increase in mortality rate is associated with a change in treatment behavior that can be represented by urologic therapies: at age 80, there is a reduction in ileoconduit usage and an increase in condom catheter use in male patients (Figure 4C2). The former may reflect shifting surgical thresholds and the latter a risk or comfort bias among treating physicians. Similarly, at age 50, there is a discontinuous decrease in prescription of the analgesic hydromorphone (Figure 4C3), suggesting a greater value placed on pain management for younger patients than for older patients. These value judgements are reflected in step changes that lead to discontinuous mortality risks at round-number ages. The model used to estimate these risk profiles does not have any algorithmic preference for round numbers, so the repeated discontinuities at round numbers suggest that these changes in risk originate from social behavior and clinical treatment.

#### Ex 4: Effects of blood urea nitrogen have flattened

Patient blood urea nitrogen (BUN) has a non-monotonic impact on mortality (Figure 4D1) with regions of rising and falling risk suggesting there are interactions between underlying biological risk and treatment effects. As expected, lower levels of BUN are associated with better survival. However, in the elevated regime, we observe several effects: from 35-60mg/dL, risk is flattened and associated with increased CRRT usage (Figure 4D2), and at 80mg/dL, risk is reduced and associated with increased treatments such as insulin (Figure 4D3) in MIMIC-IV. These associations suggest different clinical approaches to patients in these BUN ranges. Over time, as treatment protocols and clinical practice have advanced, including incoproration of BUN in multi-factor risk measures [21], the magnitude of these effects has diminished.

### Glass-box models identify opportunities to improve clinical practice

With additive models that are simultaneously interpretable and high-resolution, we can estimate the cost of round-number effects and biases, and potential improvements from refining clinical practice. Specifically, we can estimate the excess mortality attributable to biases by comparing the observed discontinuous risk to a locally-flat risk model that smooths out the observed risk to de-confound underlying biological risk from the discontinuous impact of round-number biases. For example, in the MIMIC-IV dataset, the mortality rate of patients with serum creatinine 3-6 mg/dL is elevated above the mortality rate of patients with higher serum creatinine >6 mg/dL — healthier patients exhibit higher mortality risk than less healthy patients with higher creatinine. If the risk for serum creatinine 3-6mg/dL were no worse than the observed risk for patients with serum creatinine >6 mg/dL, then we would expect a decrease of 91 deaths in the MIMIC-IV dataset of 76,540 hospitalizations with 5,653 recorded deaths (1.6%). Projecting this rate across the 500,000 ICU deaths in the United States per year [22, 23] suggests that 8049 excess deaths are attributable to the increased risk of moderately elevated serum creatinine. Thus, giving the same risk to patients with *moderately* elevated serum creatinine as is currently given to patients with *severely* elevated serum creatinine could be expected to prevent 8000 deaths in the US per year. While there may be unrecorded factors that would limit the benefit of optimized treatment, the magnitude of this effect suggests that there is an opportunity for improved treatment of patients with moderately elevated levels of creatinine to reduce their risk to that of patients with severely elevated creatinine, or at the very least, to understand the sources of these effects.

This example demonstrates the power of glass-box models to identify regions with sub-optimal clinical practice that could be refined to improve patient care. Traditionally, clinicians have not had tools to find biases and suboptimalities of care in their practice; they have had to choose between unintelligible black-box models or glass-box models such as linear and logistic regression which were less accurate and incapable of capturing the idiosyncrasies of real-world healthcare data. We propose to use modern high-resolution glass-box models to find surprising effects in observational data, and also to allow physicians and researchers to quickly identify and contextualize surprising patterns in patient outcomes and effects of clinical practice. Identifying these patterns can alert clinicians of the need to soften thresholds for particular judgments and therapeutic decisions. For example, one way these biases arise is by clinicians focusing treatment decisions on single dimensions of risk factors (i.e. one lab test at a time); if treatment decisions were instead to be guided by aggregate risk based on multivariable factors, the threshold effects would be softened. For example, recent research has shown that considering organ-organ crosstalk can improve the treatment of sepsis [24]; we propose that glass-box modeling can be used to identify and highlight surprising jumps and non-monotonicities in risk profiles that indicate opportunities for improved clinical practice, possibly including multivariable risk assessments which aggregate evidence from multiple risk factors to soften treatment decision boundaries.

## Discussion

Increasingly, AI is used to estimate and apply evidence-based statistical models of risk. In some cases these evidencebased risk models can be more accurate and more reproducible than clinician judgement [25], and offer exciting avenues for refining personalized and precision medicine [26]. But there is a fundamental challenge with data-driven models that learn to predict risk from observational medical records: patients in the training data receive informed interventions guided by patient characteristics. As a result, observed outcomes are confounded by treatments, and hence medical datasets are extremely treacherous: lurking beneath the surface of even large canonical datasets is a complex system of biomedical risk, treatment decisions, patient behavior, and real-world medical systems. Since these confounders are real-world effects, they are robustly supported by large sample sizes and confidently recapitulated by AI systems — collecting more data does not make the problem go away.

We have examined four large datasets and found that confounding by treatment effects is more the rule than the exception. This confounding is not an indictment of the model: treatment effects are real effects, so any accurate risk model trained on these datasets should recapitulate these biases. Furthermore, scale and quality of data collection do not solve this challenge: even datasets that have been collected over three decades and used by thousands of researchers to build data-driven risk models have strong treatment effects that confound data-driven risk models to estimate counter-intuitive effects. Applying such models to guide treatment decisions is risky at best, and could be harmful by incorrectly predicting low risk for patients with high underlying risk.

While these confounding effects are dangerous for blindly applying black-box models, we have shown that they also represent opportunities to improve clinical practice. Treatment effects produce characteristic statistical artifacts in risk models, including non-monotonic and/or discontinuous risk curves, and when these effects contradict biomedical causality, they can highlight regions with sub-optimal treatment distributions and potential clinical biases. We have proposed an automated system to systematize the search for these effects and demonstrated that this system can guide experts to regions of interest in which healthier patients have worse outcomes, suggesting biases in and potential improvements to clinical practice. We hope that these principles of automatically searching for discontinuities and non-monoticities in data-driven risk curves can be applied to next-generation EHR models to further optimize the practice of personalized and precision medicine.

## Materials and methods

### Datasets

#### Pneumonia

The 1989 MedisGroups Comparative Hospital Database (MCHD) pneumonia dataset [15] contains information on inpatients from 78 hospitals in 23 states in the US between July 1987 and December 1988. The MCHD contains over 250 pieces of clinical information that include patient demographic characteristics, history and physical examination findings, and laboratory and radiological results, from which 46 variables were selected [15] with known or postulated association with mortality in patients with community-acquired pneumonia. We used patient data that were collected during the first 48 hr of hospitalization. The tree-based GAMs we trained to predict mortality achieve an AUC of 0.858 on held-out patients, slightly outperforming all other models trained on this dataset [1].

#### Multiparameter Intelligent Monitoring in Intensive Care (MIMIC)

The MIMIC dataset is a high-quality collection of thousands of hospitalizations at the Beth Israel Deaconess Medical Center. This dataset has been provided in several iterations and used by thousands of researchers to estimate mortality risk models. We use three versions of this dataset: MIMIC-II [11] (24,509 hospitalizations), MIMIC-III [10] (27,349 hospitalizations), and MIMIC-IV [12] (76,540 hospitalizations). To standardize the datasets between the versions, we use only patients admitted to an ICU and select only the intersection of patient risk factors available in at least 2 datasets. The tree-based GAMs we trained achieve accuracies that are comparable or better than state-of-the-art models (Table S1).

### Methods

#### Model Estimation

We use generalized additive models (GAMs) [13] trained with boosted decision trees [27, 28] using the open-source package `InterpretML` [29]. GAMs are ideal to build glass-box models of patient risk because: (1) GAMs can be precisely decomposed into risk curves of single variables for interpretation, (2) the flexible component functions allow risk curves of any shape without any implicit preferences, (3) many treatment protocols and clinical decisions (which sum multiple sources of evidence) are inherently additive models, (4) GAMs provide the ability to edit the model [30] and reason about changes to univariable treatment protocol thresholds. We use boosted trees to train GAMs because tree-based models are scale invariant allowing features to be represented in their original natural units (including Boolean, nominal, ordinal, or continuous-valued attributes with possibly missing values) and without the bias sometimes introduced by pre-processing, and because tree-based models are better at estimating jumps and discontinuities than are spline-based GAMs, and also more likely to estimate discontinuities than smoother models such as neural nets. An important property of GAMs is that unlike simpler models such as linear and logistic regression, GAMs can estimate non-linear risk curves and are not biased as to the shape of the risk curve. The Explainable Boosting Machine (EBM) GAMs trained with `InterpretML` are particularly useful for healthcare because they use a round-robin fitting procedure that helps ensure that all hidden effects are observable in the estimated model. We benchmark the model against several baselines (Table S1), including several versions of `XGBoost` and fully-connected neural networks to verify that there would not be a gain in accuracy with interaction effects.

#### Automated Search

To automate the search for discontinuities and non-monotonicities, we define two statistical tests on the likelihood of the shape of a component (Figure S2). For each component function *f*_*r*_ estimated by the tree-based GAM, we have a set of thresholds *t*_*r*_ and associated probability densities *y*_*r*_. We evaluate the likelihood of a discontinuity at threshold *t*_*r,j*_ by comparing the probability of observed outcomes under the discontinuous threshold-based component function *f*_*r*_ against the probability of observed outcomes under a linearized version of *f*_*r*_ (Figure S2B). We evaluate the likelihood of a non-monotonicity at threshold *t* in component *r* by applying changepoint detection [14] to the sign of non-zero slopes of the component function (Figure S2C).

## Data Availability

No new data are presented in the manuscript. Code to reproduce analyses are available at the following link.

https://github.com/blengerich/DeathByRoundNumbers

## Code availability

`Python` notebooks reproducing these analyses, and utilities for automated search and plotting of discontinuities and non-monotonicites are available at: github.com/blengerich/DeathByRoundNumbers.

## Data availability

All MIMIC datasets analyzed in this study are distributed freely under the PhysioNet platform [31]. The Pneumonia dataset analyzed in this study as a motivating example is available from the corresponding author on reasonable request. Intermediate data generated for analyses are available in the `Python` notebooks linked above.

## Supplemental Information for

### S1 Simulation Study

To simulate the effects of confounding by treatment effects, we modeled intrinsic risk of a risk factor as *P* (*Y*|*X*) = *σ*(*f* (*x*)) = 2 * (*x* − 0.5)^2^, where *σ* is the logistic function. This simulates a healthy region surrounded by two unhealthy regions, with risk continuously increasing as the biomarker value moves away from the healthy range. We sample *x* ∈ [0.35, 1.0], and apply *f* (*x*) to generate an empirical set of risks *R*. We model four different types of treatments to generate treated risks *g*(*x*):

- **Treatment Flattens Risk**: *g*(*x*) = *P*_50_(*R*)
- **Treatment Caps Risk**: *g*(*x*) = min(*f* (*x*), *P*_50_(*R*))
- **Treatment Reduces Biomarker**: *g*(*x*) = *f* (*x* − 0.2)
- **Treatment Has Constant Benefit**: *g*(*x*) = *f* (*x*) − *P*_75_(*R*)

We simulate two versions of treatment probabilities:

- **Strict Adherence**: 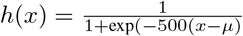
- **Loose Guidance**: 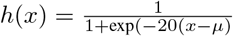

The threshold *μ* of the treatment protocol is determined by whether the threshold should be “optimal” (*μ* = *μ*_0_ = argmin_*x*_ *f* (*x*) *< g*(*x*)), “too low” (*μ* = *μ*_0_ − 0.2), or “too high” (*μ* = *μ*_0_ + 0.2). These treatment probabilities are visualized in Figure S1.

**Figure S1:**
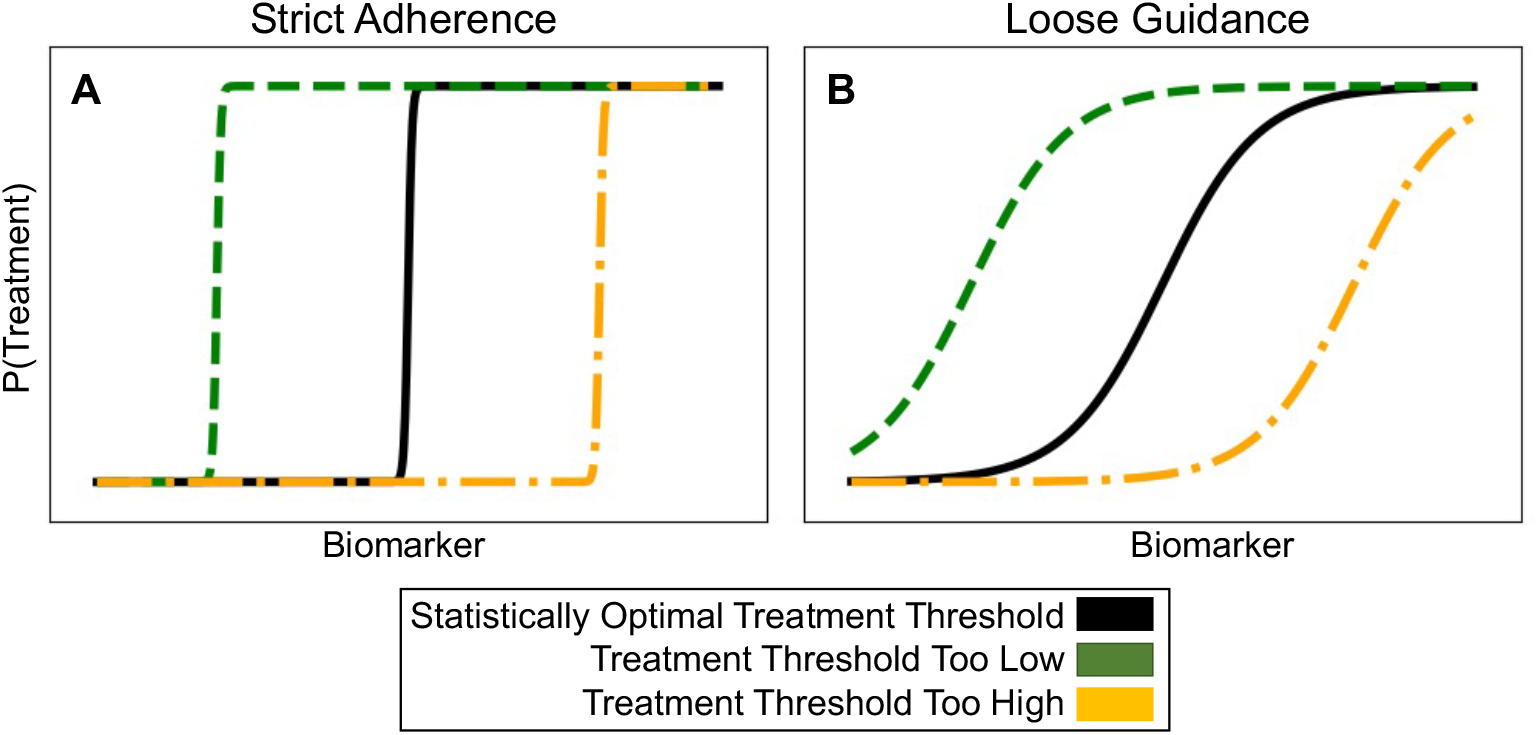
Simulated treatment probabilities. **(A)** Treatment probabilities which follow strict adherence to a threshold-based protocol. **(B)** Treatment probabilities which treat the threshold-based protocol as loose guidance.

Given a treatment risk function and a treatment protocol function, we simulate observed population risk by *q*(*x*) = *f* (*x*)(1 − *h*(*x*)) + *g*(*x*)*h*(*x*). Effects of hyperparameters do not change the fundamental characteristic shapes; code to reproduce these results are available at github.com/blengerich/DeathByRoundNumbers.

### S2 Automated detection of discontinuities and non-monotonicities

As described in the Methods section of the main text, we used a generalized additive model (GAM) to estimate mortality risk from observational data (Figure S2). The only change we make to the default hyperparameters is to increase the number of rounds of bootstrap sampling to 100, following the convention suggested by the algorithm designers [1]. `Python` notebooks reproducing these analyses, and utilities for automated search and plotting of discontinuities and non-monotonicites are available at: github.com/blengerich/DeathByRoundNumbers.

#### Automated detection of discontinuities

Let *r* be the component of interest, with component function *f*_*r*_ estimated by the tree-based GAM defining a set of thresholds *t*_*r*_ and associated outcome odds *y*_*r*_. We construct a locally-linear version of *f*_*r*_:

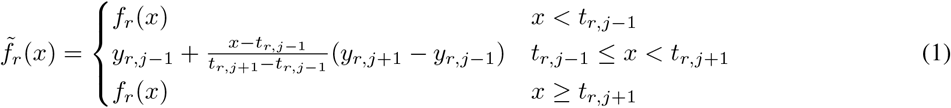

To evaluate a discontinuity at threshold *t*_*r,j*_, we define a test statistic *T*_*d*_ which compares the log-likelihood of the discontinuous component against the log-likelihood of the linearized component:

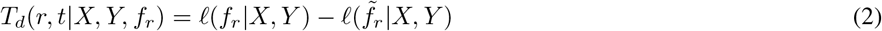

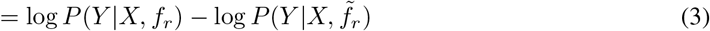

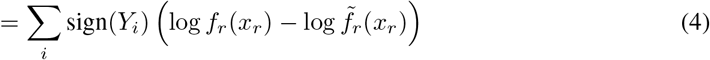

where *i* indexes observed samples and the final equality holds by additivity of component functions in the GAM. This test statistic estimates the likelihood of a discontinuity at each threshold; bootstrap provides confidence intervals.

#### Automated detection of non-monotonicities

Let *r* be the component of interest, with component function *f*_*r*_ estimated by the tree-based GAM defining a set of thresholds *t*_*r*_ and associated outcome odds *y*_*r*_. We calculate the slopes *s*_*r*_:

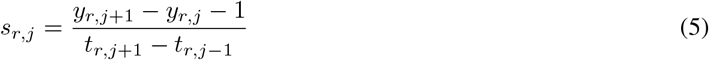

and perform changepoint detection [14] on the signs of *s*_*r*_. This statistic estimates the likelihood of a change in the direction of the component function at each threshold; bootstrap provides confidence intervals. Finally, we restrict the search to only concave non-monotonicities because convex risk curves are usually explainable as healthy states.

**Figure S2:**
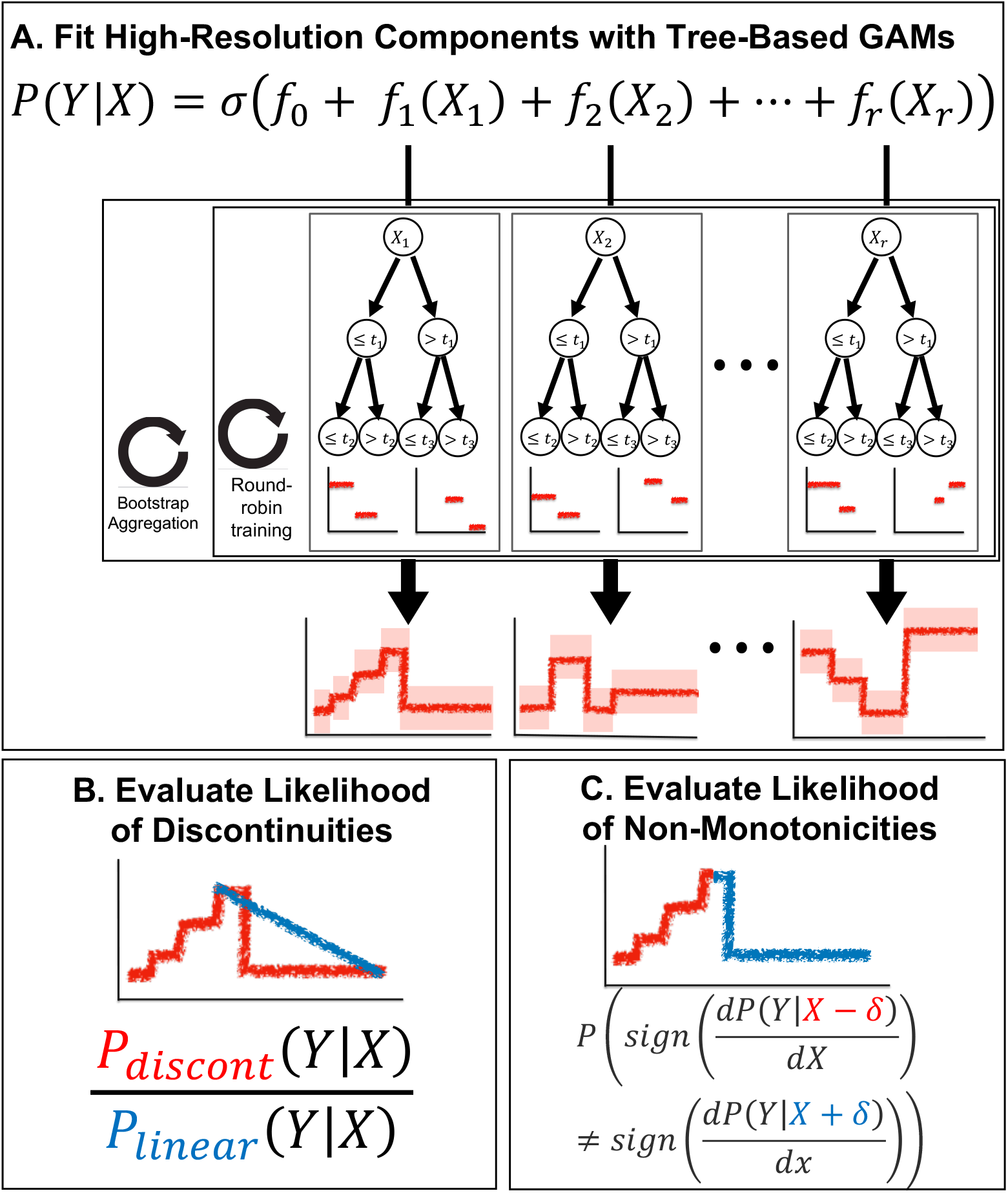
Summary of our approach to estimate outcome probabilities and automate search for statistical artifacts of clinical practice. **(A)** Sketch of the tree-based GAM trained by `InterpretML`, which models the logit probability of outcome *Y* as an additive function of individual features of *X*. This model uses round-robin training to split information between individual component functions and bootstrap aggregation for confidence intervals. **(B)** For each component function and each threshold therein, we evaluate the likelihood of a discontinuity by comparing the probability of observed data under the discontinuous component function against a locally-linear version of the component function. **(C)** For each component function and each threshold therein, we evaluate the likelihood of a non-monotonicity by appyling changepoint detection to the nonzero slopes of the signs of the component function.

### S3 Accuracy of GAM

In general we find that GAMs trained with EBMs provide a sweet spot for modeling healthcare data because the models are complex enough to be very accurate and represent high-resolution details such as discontinuities while still remaining fully interpretable. Moreover, because the models are restricted in complexity, they are much less likely to learn spurious effects that do not generalize to held-out examples. Finally, `InterpretML` uses bootstrap resampling [32] to reduce variance and make the learned models easier to interpret, and also to estimate confidence intervals that help distinguish true from spurious detail in the learned response curves. We benchmark the model against several baselines (Table S1) to examine the performance of GAMs with EBMs. As baselines, we include 3 versions of XGBoost [33] which differ in the max depth (XGB-1 creates trees of depth 1, XGB-2 creates trees of depth 2, and XGB-3 creates trees of depth 3). As a result, XGB-1 is a GAM which can be directly compared against the EBM GAM, and the performance of the deeper XGBoost versions can be compared to evaluate the utility of permitting interaction effects in the model. Finally, we also benchmark against fully-connected neural networks (MLP). In all four of these datasets, the EBM performs as well or better than the baseline methods by AUROC.

**Table S1:** Accuracy metrics of the predictions made by EBM and other models on the held-out test set of patients demonstrate that EBM is near or better-than comparable general-purpose models on these datasets. Values are the (mean*pmstd*) over 5 experimental runs.

| Dataset | Model | AUROC | AUPRC | F1-Score |
| --- | --- | --- | --- | --- |
| Pneumonia | XGB-1 | $0.86 \pm 0.01$ | $0.47 \pm 0.01$ | $0.34 \pm 0.01$ |
| | XGB-2 | $0.84 \pm 0.01$ | $0.43 \pm 0.02$ | $0.34 \pm 0.01$ |
| | XGB-3 | $0.80 \pm 0.04$ | $0.39 \pm 0.02$ | $0.35 \pm 0.03$ |
| | MLP | $0.75 \pm 0.05$ | $0.29 \pm 0.02$ | $0.22 \pm 0.03$ |
|  | EBM | **$0.88 \pm 0.03$** | **$0.53 \pm 0.02$** | **$0.37 \pm 0.02$** |
| MIMIC-II | XGB-1 | $0.78 \pm 0.02$ | $0.35 \pm 0.02$ | $0.18 \pm 0.01$ |
| | XGB-2 | $0.78 \pm 0.02$ | $0.35 \pm 0.02$ | $0.18 \pm 0.02$ |
| | XGB-3 | $0.77 \pm 0.05$ | $0.35 \pm 0.02$ | **$0.28 \pm 0.02$** |
| | MLP | $0.74 \pm 0.04$ | $0.30 \pm 0.03$ | $0.07 \pm 0.02$ |
| | EBM | **$0.79 \pm 0.02$** | **$0.38 \pm 0.03$** | $0.17 \pm 0.02$ |
| MIMIC-III | XGB-1 | $0.73 \pm 0.02$ | $0.26 \pm 0.01$ | $0.07 \pm 0.00$ |
| | XGB-2 | $0.72 \pm 0.04$ | $0.23 \pm 0.02$ | $0.11 \pm 0.00$ |
| | XGB-3 | $0.69 \pm 0.05$ | $0.22 \pm 0.05$ | **$0.14 \pm 0.01$** |
| | MLP | $0.68 \pm 0.08$ | $0.24 \pm 0.03$ | $0.03 \pm 0.01$ |
| | EBM | **$0.76 \pm 0.01$** | **$0.31 \pm 0.01$** | $0.06 \pm 0.01$ |
| MIMIC-IV | XGB-1 | $0.74 \pm 0.01$ | $0.24 \pm 0.01$ | $0.10 \pm 0.03$ |
| | XGB-2 | **$0.75 \pm 0.03$** | **$0.27 \pm 0.03$** | $0.17 \pm 0.03$ |
| | XGB-3 | $0.73 \pm 0.05$ | $0.25 \pm 0.03$ | **$0.20 \pm 0.02$** |
| | MLP | $0.73 \pm 0.02$ | $0.25 \pm 0.02$ | $0.13 \pm 0.01$ |
| | EBM | **$0.75 \pm 0.03$** | $0.26 \pm 0.01$ | $0.08 \pm 0.01$ |

### S4 Extended analysis of pneumonia dataset

**Figure S3:**
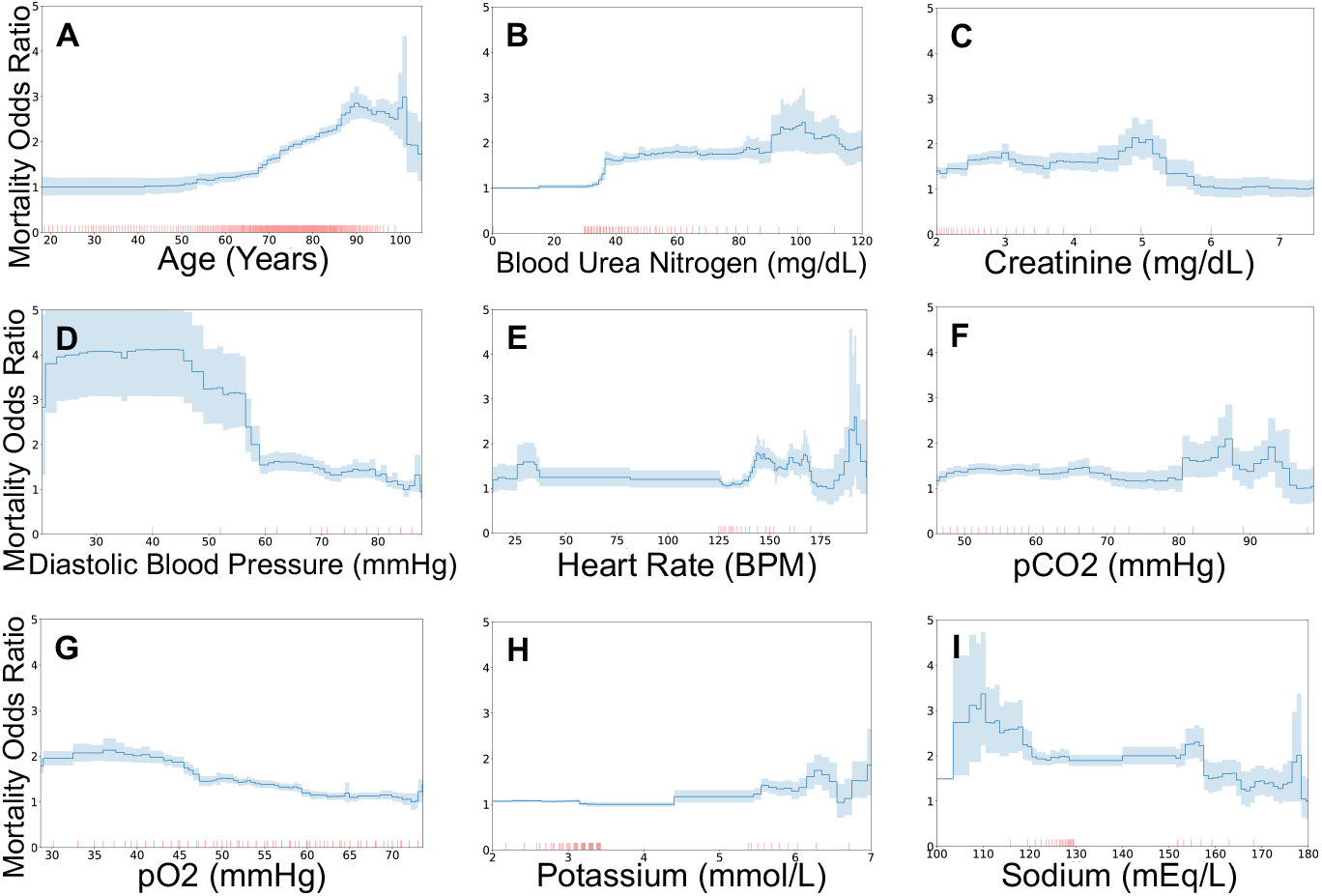
Mortality risks of patients hospitalized with pneumonia display several treatment effects. In all plots, we display the odds ratio after correcting for all other patient risk factors. Shaded regions indicate 95% confidence intervals, and each red tick mark along the horizontal axis denotes 10 patients. In this dataset, values in the reference (healthy) range were not recorded. **(A)** Mortality risk rises gradually with increased age, with a change in slope at age 65, a rapid rise in risk approaching age 90, a counter-causal plateau in risk for ages 90-100, and a discontinuous drop in risk for at age 100. **(B)** Mortality risk is low for blood urea nitrogen below 30 mg/dL and then rises rapidly to a plateau starting at 35 mg/dL. **(C)** Mortality risk is lowest for several elevated levels of serum creatinine, which indicates kidney dysfunction. **(D)** Mortality risk rises slowly as diastolic blood pressure decreases, with a rapid rise at 60 mmHg followed by a plateau in risk. This pattern would agree with treatment decisions based on a blood pressure of 60mmHg, and the statistically-optimal threshold may be lower than 60mmHg. **(E)** Mortality risk rises as heart rate increases above 125 BPM, but plateaus from 140-170 BPM, and drops at 170 BPM. This risk profile indicates clinical decisions that change based on tachycardia levels of 140 and 170 BPM and a potential improvement to clinical practice to benefit patients with heart rate 140-170 BPM. **(F)** At 80 mmHg pCO2, there is a jump in risk followed by a plateau and slow decline. This suggests treatment decisions could be enhanced to flatten this risk curve. **(G)** Mortality risk changes smoothly with pO2, with a gradual increase in risk as pO2 decreases. This indicates well-tuned treatment decisions that do not exhibit strong potential for further refinement by changing thresholds. **(H)** Mortality risk rises gradually for elevated levels of serum potassium, with a small rise in risk at 5.5mmol/L followed by a plateau in risk. This suggests a treatment threshold at 5.5 mmol/L. **(I)** As expected, mortality risk for patients with hyponatremia <120 mEq/L is elevated. However, mortality risk for patients with severe hypernatremia >155 mEq/L is reduced, suggesting that clinical practice could be refined to benefit patients with moderate hypernatremia.

### S5 Extended Analysis of MIMIC

**Figure S4:**
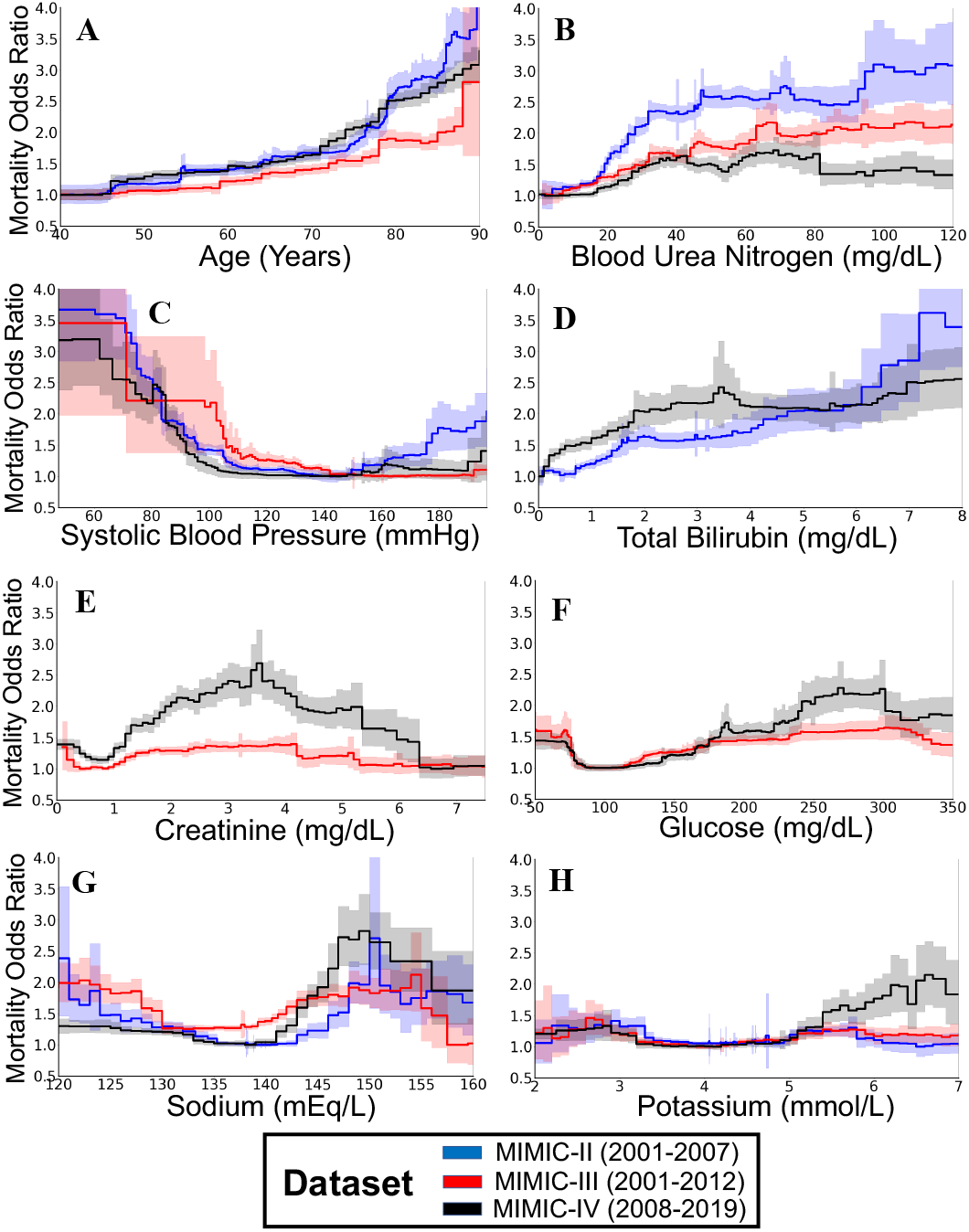
The effects of clinical decisions are visible in mortality risk curves for patients in intensive care units, and have changed over time. In all plots, we show the risk associated with each risk factor after correcting for all other observable factors of patient risk using a generalized additive model (with 95% confidence intervals shaded). The three datasets span three decades of intensive care at a single hospital system. The impacts of creatinine, sodium, age, and blood urea nitrogen are studied in detail in the main text. **(A-D)** Biomarkers that display discontinuous impacts of treatment effects, including round-number ages, blood urea nitrogen at 45mg/dL, systolic blood pressure below 100 mmHg, and Bilirubin above 2mg/dL. **(E-H)** Biomarkers which display counter-causal impacts of treatment effects, including elevated creatinine above 6mg/dL corresponding to reduced mortality risk, elevated glucose above 250 or 300 mg/dL corresponding to reduced mortality risk, hypernatremia above 150 mEq/L corresponding to reduced mortality risk (and in the oldest dataset, severe hyponatremia below 120 mEq/L also corresponded to reduced mortality risk), and elevated potassium (above 6 mmol/L) corresponding to increased mortality risk only in the newest dataset.

